# Quality and Safety profiles of AI-Generated vs Clinician-Generated Handoffs in Hospital Medicine

**DOI:** 10.64898/2026.06.05.26354946

**Authors:** Kaustav P. Shah, Subha Airan Javia, Thomas Savage, Eric Bressman

## Abstract

**Introduction:** End-of-rotation handoffs are critical for patient safety but add to documentation burden for hosptialists. Generative artificial intelligence (AI) may help automate handoff creation using electronic health record data, but its impact on quality and safety is unclear.

**Methods:** We developed an AI handoff tool with a large language model using clinical notes as input and conducted a retrospective evaluation comparing AI-generated and clinician-authored handoffs. Handoffs were assessed across domains of quality and safety through a structured review.

**Results:** Quality ratings were similar between AI and human handoffs (3.7 vs. 3.5, p=0.57). AI-generated handoffs were rated higher for organization (4.4 vs. 4.1, p=0.05) and completeness (4.1 vs. 3.6, p=0.01), but lower for conciseness (3.7 vs. 4.1, p=0.03) and accuracy (4.1 vs. 4.4, p=0.03). Error rates were comparable (0.3/handoff in both groups); however, AI-generated handoffs included inaccuracies (9% of AI errors) and hallucinations (1% of AI errors), while clinician-authored handoffs contained only omissions.

**Conclusion:** Human and AI handoffs have differing error profiles and tradeoffs between completeness and conciseness. Prospective evaluation in clinical workflows is underway.

## Introduction

End-of-rotation handoffs (or sign-outs), in which care is transitioned between hospitalists at the end of a service block, are important junctures for patient safety [1]. Prior studies have demonstrated an increased risk of adverse events around this time [2]. Guidelines recommend that handoffs clearly label anticipated events and action items [3], yet in practice, critical clinical information is often omitted, leading to communication failures that may negatively impact patient outcomes [4, 5]. In addition, written handoff, while likely superior to verbal-only communication [6], represents an additional documentation task for clinicians – on top of routine progress notes, discharge summaries, and coding queries – contributing to the growing burden of electronic documentation and clinician burnout [7].

Generative artificial intelligence (AI) tools, including large language models (LLMs), are capable of processing large volumes of unstructured clinical data and generating coherent text [8]. Early studies have evaluated LLMs for drafting clinical documentation, such as discharge summaries, with performance comparable to that of physicians [9]. However, little work has examined the use of LLMs for handoff-related tasks, which differ from other forms of clinical documentation in their intended audience, style, and function. In this study, we developed an AI handoff tool using a commercially available LLM and compared the quality and safety profiles of AI and human-generated handoffs.

## Methods

We developed an AI-handoff tool using a secure instance of an LLM (OpenAI, GPT-o1[10]). The tool was embedded within Carelign, a locally developed, team collaboration platform used for handoffs at the University of Pennsylvania Health System that interfaces with the electronic health record (EHR) [11]. To optimize model output, we conducted iterative prompt engineering focusing on two components: 1) the prompt instructions and 2) the clinical notes provided as contextual input to the LLM. The final prompt configuration was selected through consensus review by the study team. The clinical notes provided as contextual input included: 1) the most recent hospital medicine progress note (or admission note if no subsequent progress note was available) and 2) the most recent notes from any specialty consultants signed within the previous 72 hours. The final prompt text is provided in the Appendix.

To assess the quality and safety of the tool prior to clinical deployment, we developed a structured review process comparing AI-generated and clinician-generated handoffs. Nine hospitalists were recruited and asked to provide previously composed handoffs for ten patients they had cared for in the prior year. The AI handoff tool was then used by the study team to retrospectively generate handoffs for these patients using clinical notes from the same time point, resulting in paired human and AI handoffs. AI-generated handoffs were not edited prior to review and were generated in August 2025.

Each hospitalist reviewer was assigned to evaluate five human-AI handoff pairs (10 total handoffs) for patients with whom they had no prior clinical involvement along with five AI-generated handoffs for patients they had previously cared for, resulting in an expected total of 15 reviews per reviewer. Reviewers were blinded to whether the handoff was generated by human or AI and were encouraged to access the EHR as needed to verify information for patients they had not cared for.

The structured review assessed overall quality, completeness, organization, conciseness, and accuracy using 5-point Likert scales. Reviewers were also asked to identify errors in the handoffs; estimate their potential for harm and likelihood of impacting the patient’s care using 5-point Likert scales; and classify them as omissions (key information missing from handoff), hallucinations (information generated in the handoff that did not exist in the source record), or inaccuracies (information incorrectly reported in the handoff). Reviewers additionally provided qualitative feedback on handoff quality through free-text comments. The rubric was modeled off existing frameworks and published reports to assess quality and safety for LLMs in medical contexts [9, 12].

Two-sided T-tests were used to compare AI and human-generated handoffs across quality and safety domains measured on Likert scales. Fisher’s exact test was used to compare error and omission rates between the AI and human-generated groups. The study was approved by the Institutional Review Board at the University of Pennsylvania.

## Results

A total of 133 handoffs were reviewed, including 88 AI-generated and 45 human-generated handoffs (Table 1). Of the 135 handoffs assigned for review, two were not completed due to reviewer error.

**Table 1:** Comparison of Human and AI handoffs across domains of documentation quality.

| Domain | AI handoff score<br>Mean (SD) | Human handoff score<br>(mean, SD) | P value |
| --- | --- | --- | --- |
| Quality |  |  |  |
| Overall quality | 3.7 (1.0) | 3.5 (1.3) | 0.57 |
| Conciseness | 3.7 (1.3) | 4.1 (0.9) | 0.03 |
| Organization | 4.4 (0.7) | 4.1 (0.9) | 0.05 |
| Completeness | 4.1 (0.9) | 3.6 (1.2) | 0.01 |
| Accuracy | 4.1 (0.9) | 4.4 (0.7) | 0.03 |

Overall quality ratings were similar between AI and human handoffs (3.7 vs. 3.5, p=0.57). AI handoffs were rated higher for organization (4.4 vs. 4.1, p=0.05) and completeness (4.1 vs. 3.6, p =0.01), but lower for conciseness (3.7 vs. 4.1, p=0.03) and accuracy (4.1 vs. 4.4, p=0.03). Reviewer quotes focused on a trade-off between length and detail of information in the handoff.

Across the 88 AI-generated handoffs, reviewers identified 28 errors (0.3 errors per handoff), including 1 hallucination, 9 inaccuracies, and 18 omissions (Table 2). Among the 45 human-generated handoffs, 13 errors were identified (0.3 errors/handoff), all of which were omissions. The difference in proportion of omissions was statistically significant between the two groups (p=0.02). Ratings of potential harm (AI: 1.9, human: 1.9) and likelihood that an error would impact patient care (AI: 2.4, human: 2.3) were similar between groups. Reviewer free text comments supported error labels, including concerns about misrepresentation of discharge plans and consultant recommendations, with representative quotes included in Table 2.

**Table 2:** Comparison of errors across Human and AI handoffs.

| Domain | AI handoff, Mean (SD), n = 28 | Human handoff Mean (D), n = 13 | P value | Representative Quotes – AI handoffs | Representative Quotes – Human handoffs |
| --- | --- | --- | --- | --- | --- |
| Potential harm of error | 1.9 (0.5) | 1.9 (0.5) | 0.97 | “Left out that the patient had a PEG previously (important context given his current swallowing issues)” | “Should have had info re. agitation/baseline mental status and that he was on a 1:1 and why.” |
| Likelihood of error impacting patient’s care | 2.4 (1.0) | 2.3 (1.0) | 0.89 | “Reason for admission is incorrect. She was admitted following ground-level fall while on anticoagulation. She remained admitted for OUD management.” | “missing detail about dc meds and exact home care plan (palliative vs hospice), this info would be particularly useful given lack of provider continuity and plan for weekend discharge so also no CM/SW continuity” |
| Errors labeled as omission n (% of total) | 18 (64) | 13 (100) | 0.02 | “They forgot about the cat not having a caregiver - important bc pt might not go to snf bc of that” | “This seems to be a human generated one. It is unbelievably short. Like no mention of how bad the AKI was, shock = to what degree like pressors for many days or just IV fluids, not sure which cultures are growing Strep mitis unless it's only the blood.” |
| Errors labeled as inaccuracy n (% of total) | 9 (32) | 0 (0) |  | “At the time of this handoff, dispo was pt recommended to acute rehab (not home or SNF, as noted)” | N/A |
| Errors labeled as hallucination n (% of total) | 1 (4) | 0 (0) |  | “Acting like we knew she's follow with *oncology* after discharge and that onc wasn't going to offer until they reviewed her pathology (which yes - makes sense, but they had not confirmed this with us yet)” | N/A |

## Discussion

We developed an AI-based tool to draft hospitalists’ end-of-rotation handoffs using a secure instance of a commercially available LLM. Overall quality ratings were similar between AI and human-generated handoffs. Human handoffs were rated as more concise and accurate, whereas AI handoffs were rated as more organized and complete. These differences likely reflect tradeoffs inherent in the prompt engineering process, and future iterations of this type of tool may allow for customization of output to better align with individual user preferences. These findings align with emerging studies demonstrating that LLMs can assist with clinical documentation tasks while introducing distinct error profiles that require careful workflow integration.

Higher ratings for conciseness and accuracy in human handoffs likely reflect clinicians’ ability to apply clinical judgement and highlight the most pertinent information for the receiving provider. In contrast, the higher organization scores for AI handoffs likely reflect the use of a structured prompt and predefined output template. AI handoffs were also rated as more complete, which is not unexpected given that the model had access to multiple recent clinical notes and is not subject to fatigue in data review or handoff composition. Together, these findings suggest a complementary approach in which an AI-generated draft provides a comprehensive and structured starting point that clinicians can further refine using their clinical judgement as a future workflow that should be tested.

Although overall error rates were similar, the types of errors differed between groups. Errors in human handoffs contained exclusively omissions, a pattern consistent with prior studies demonstrating that missing information is a common source of communication failures during handoffs and may contribute to adverse patient outcomes [5]. In contrast, more than a third of AI errors were hallucinations or inaccuracies, a known limitation of LLMs [13]. In our study, the AI handoff tool occasionally reported discharge plans with unfounded certainty when documentation indicated that plans were still under deliberation. Reviewers rated the potential magnitude of harm and the likelihood of an error reaching a patient as equivalent even though the salience to a clinician of misrepresentation may be higher than omitting information. AI handoffs in this study were reviewed in their unedited form; in a future clinical workflow, a clinician would ideally review and edit such errors before sharing the handoff with a colleague. Still, a “human-in-the-loop” strategy may not fully mitigate risks associated with AI-generated errors[14] as automation bias may result in suboptimal review and editing of AI outputs without appropriate training and human factors design [15].

This study has several limitations. First, the evaluation was retrospective and relied on reviewers’ subjective assessments of handoff quality and safety. Each reviewer’s observation was treated independently, and reviewers could use EHR as they deemed appropriate to verify handoff information. Second, the study was conducted within a single hospital medicine group in one health system that does not use a standardized handoff template, which may limit generalizability. The prompt for the AI handoff specifies an output style partially explain higher scores for AI in the organization domain. Third, the tool was built using an earlier generation LLM (GPT-o1) which is no longer commonly used. Fourth, despite blinding, many reviewers commented on their ability to identify handoffs as AI or human-generated based on the style and patterns in the text. Despite these limitations, to our knowledge this study represents the first systematic evaluation of large language model-generated end-of-rotation handoffs compared with clinician-authored handoffs, using blinded reviewers and structured assessment of quality and safety domains.

Future work should evaluate the impact of AI-generated handoffs in real-world clinical workflows and incorporate strategies including tool use that can actively scan the EHR rather than relying on predefined set of clinical notes. A pragmatic randomized trial is underway to prospectively evaluate the AI handoff tool in hospital medicine workflows to determine if it can improve documentation time and provider well-being.

## Data Availability

Data contains protected patient specific information that cannot be shared broadly, summary data on review ratings may be shared on reasonable request to the authors

## Disclosures

The views expressed are those of the authors and do not necessarily reflect the position or policy of the VA or the US government.

Thomas Savage currently works for Doctronic, an AI care delivery company. Doctronic has no connection to this work.

## Contribution statements

KS: conceptualization, funding acquisition, data curation, formal analysis, writing – original draft, methodology

TS: conceptualization, investigation, methodology, writing – review and editing

SAJ: Conceptualization, Funding acquisition, Methodology, Software, Supervision, Writing - review & editing

EB: Conceptualization, Formal analysis, Funding acquisition, Methodology, Supervision, Writing - review & editing

